# Healthcare workers’ worries and Monkeypox vaccine advocacy during the first month of the WHO Monkeypox alert: Cross-sectional survey in Saudi Arabia

**DOI:** 10.1101/2022.08.02.22278317

**Authors:** Fadi Aljamaan, Shuliweeh Alenezi, Khalid Alhasan, Basema Saddik, Ali Alhaboob, Esraa S. Altawil, Fatimah Alshahrani, Abdulkarim Alrabiaah, Ali Alaraj, Khaled Alkriadees, Yousef Alshammari, Homood Alharbi, Amr Jamal, Rabih Halwani, Fahad AlZamil, Sarah Al-Subaie, Mazin Barry, Ziad A Memish, Jaffar A. Al-Tawfiq, Mohamad-Hani Temsah

## Abstract

**Background:** Monkeypox virus re-surged in May 2022 as a new potential global health threat with outbreaks bursting in multiple countries across different continents. This study was conducted during the first month of the WHO announcement to assess the healthcare workers (HCWs) within Saudi Arabia, exploring their perception, worries, and vaccine acceptance for Monkeypox in-line with the resolving COVID-19 pandemic.

**Methods:** A national cross-sectional survey was conducted between May 27 and June 10, 2022, in Saudi Arabia. Data were collected on the sociodemographic and job-related characteristics, COVID-19 infection status, HCWs’ worry levels of Monkeypox compared to COVID-19 and its sources, their perceptions, awareness, and HCWs’ Monkeypox vaccination advocacy.

**Results:** Among the 1130 HCWs who completed survey, 41.6% already developed COVID-19. Still, 56.5% were more worried from COVID-19 compared to Monkeypox, while the rest were more worried of Monkeypox disease. The main reason for their worry among 68.8% of the participants was development of another worldwide pandemic post COVID-19, followed by their worry of acquiring the infection themselves or their families (49.6%). Most HCWs (60%) rated their self-awareness of Monkeypox disease as moderate to high.

Males and those who previously developed COVID-19 were significantly less likely to worry about Monkeypox. The worry about Monkeypox developing into a pandemic and the perception of Monkeypox being a severe disease correlated significantly positively with the odds of high worry from the disease.

Regarding participants’ advocacy for HCWs’ vaccination against Monkeypox disease, those who developed COVID-19 previously and those who supported application of tighter infection control measures compared to the current ones to combat the disease were significantly predicted to agree for vaccination. 74.2% of the surveyed HCWs perceived that they need to read more about the Monkeypox disease after the survey.

**Conclusion:** During the first month of the WHO’s Monkeypox international alert, about half of HCWs in this study were more worried about Monkeypox disease as compared to COVID-19, and its possible progression into another pandemic. In addition, the majority were in favor of applying tighter infection prevention measures to combat the disease. The current study highlights areas needed for healthcare administrative about the HCWs’ perceptions and readiness for Monkeypox especially in the event of any occurrence of local or international pandemic.

## Introduction

The recent reemergence of the Monkeypox virus (MOXV) had caused a significant worry of the potential development of another pandemic on the shadow of the existing COVID-19 pandemic. (MOXV) is zoonotic in origin with multiple reservoirs. (MOXV) refers to the first isolation from captive monkeys shipped to the Netherlands from Africa in 1958 and later was identified for the first time in human in 1970[1]. The recent multi-countries outbreak of (MOXV) with absence of travel to endemic areas is certainly a cause of concern[1,2]. This concern was exemplified by scientists around the globe[3]. The World Health Organization (WHO) called for an international meeting to discuss the importance of (MOXV), which concluded that the event did not constitute a Public Health Emergency of International Concern (PHEIC). However, the emergence of (MOXV) still adds to the current burden of anxiety of HCWs as well as the public[4]. A recent questionnaire revealed that about 62% of the general population were more concerned about (MOXV) in comparison to COVID-19[5].

The continued COVID-19 pandemic is associated with stress among HCWs as well as increased workload and anxiety[6]. One of the approaches to control the current outbreak of (MOXV) is vaccination. Such strategy is being utilized in certain countries such as the US, Canada and Europe[7]. However, such a strategy is associated with challenges and unknowns[7]. The occurrence of 521 cases of (MOXV) in Germany showed a median age of 38 years and all cases were men[8]. Similarly, the reported cases from Spain showed predominance of men who had sex with men[9]. Other studies had looked at the global estimates of (MOXV) cases across several nations[10,11]. On the other hand, the COVID-19 pandemic had facilitated many countries around the globe to develop rapid testing, isolation, and management[10]. Some countries had deployed Research Electronic Data Capture (REDCap) design and programmers and infection control activities in response to (MOXV) outbreaks globally[12]. Amid these unknowns and the need to develop a global strategy to combat the emergence of (MOXV), we developed rapidly a survey directed towards HCWs in Saudi Arabia in order to examine their worries and Monkeypox vaccine advocacy during the first month of the WHO Monkeypox alert.

## Method

### Data Collection

An online survey of healthcare workers in the Kingdom of Saudi Arabia (KSA) was conducted over ten days (from May 27 to June 5, 2022). Participants were invited by convenience sampling techniques through various social media platforms (Twitter and WhatsApp groups) and email lists. Participants were asked to complete the online survey through the SurveyMonkey^©^ platform, with each response allowed once from each unique IP address to ensure single entries. The first page of the survey included consent of participation, explained the research objectives and assured confidentiality.

The survey tool was adopted from our published research on COVID-19 with modifications related to the new Monkeypox outbreak [13-17]. The final survey version was piloted among ten HCWs for clarity and consistency. Modifications were implemented based on the experts’ recommendations. The questionnaire took eight minutes to complete. The research team approved the final version of the survey for language accuracy, clarity, and content validity.

Variables surveyed included HCWs’ sociodemographic and job-related characteristics, levels of worries from Monkeypox compared to COVID-19 and sources of worries, previous COVID-19 infection status, advocacy for Monkeypox disease vaccination. Also, we assessed their compliance with infection prevention precautionary measures against COVID-19 and their support for tighter measure in line with the (MOXV) outbreaks. Finally, their Generalized Anxiety Disorder (GAD7) score[18,19] (GAD7) which is a self-reported, 7-item validated scale was assessed.

#### 1.1 Ethical Approval

Ethical approval was granted by the institutional review board (IRB) at King Saud University (22/0416/IRB) before the collection of data.

#### 1.2 Statistical analysis

Means and standard deviations were used to describe continuous variables and frequencies and percentages for categorically measured variables. The histogram and the Kolmogorov-Smirnove test were applied to test the assumption of normality, and the Levene’s test was used to test the homogeneity of variance statistical assumption. Cronbach’s alpha test was used to assess the internal consistency of the measured questionnaires. The Multivariate Binary Logistic Regression Analysis was used to assess variables independently correlating with HCWs’ worry from (MOXV), their advocacy for HCWs’ vaccination against it, and their perception of need to apply tighter infection prevention practices (IPC) of the current in order to combat the (MOXV) outbreaks. The association between predictors with the outcome dependent variables in the multivariate Logistic Binary regression analysis was expressed with adjusted Odds Ratio (OR) with their associated 95% confidence intervals. The SPSS IBM statistical analysis program was used for statistical data analysis. The statistical Alpha significance level was considered at 0.050 level.

## Results

1130 HCWs residing in Saudi Arabia participated in the study. Table-1 displays the sociodemographic and professional characteristics. The mean age was (37.1, SD= 9.69 years). Of the participants, 62.7% were females 62.7%, 66.2% were married 66.2%, and 57% were expatriate. The details of their household size and monthly income (HHI) are displayed in Table 1.

**Table-1:** Participants’’ baseline sociodemographic and professional characteristics. N= 1130.

| Characteristic | Frequency | Percentage |
| --- | --- | --- |
| **Sex** |  |  |
| Female | 708 | 62.7 |
| Male | 422 | 37.3 |
| **Age group** |  |  |
| 20—30 years | 321 | 28.5 |
| 31—40 years | 450 | 39.9 |
| 41—50 years | 227 | 20.1 |
| ≥ 51—60 years | 129 | 11.5 |
| **Marital state** |  |  |
| Never married | 381 | 33.8 |
| Married | 746 | 66.2 |
| **Nationality** |  |  |
| Saudi | 486 | 43 |
| Expatriate | 644 | 57 |
| **Households (family) size —members** |  |  |
| Prefer not to answer | 122 | 10.8 |
| 1—3 members | 446 | 39.6 |
| 4—6 persons | 408 | 36.2 |
| 7—10 persons or more | 151 | 13.4 |
| **Households’ monthly income (Saudi Riyals)** |  |  |
| Prefer not to answer | 354 | 31.4 |
| Less than 10000 SR | 257 | 22.8 |
| 10001—15000 SR | 118 | 10.5 |
| 15001—20000 SR | 120 | 10.6 |
| > 20000 SR | 278 | 24.7 |
| **Working hospital type** |  |  |
| Primary healthcare center | 225 | 20 |
| Secondary—care hospital | 202 | 17.9 |
| Tertiary hospital | 700 | 62.1 |
| **Clinical Role** |  |  |
| Medical student | 80 | 7.1 |
| Technicians/Lab workers and Pharmacists | 132 | 11.7 |
| Nurses | 470 | 41.7 |
| Physicians | 445 | 39.5 |
| **Hospital Working area/ covering service** |  |  |
| Pharmacy and laboratory | 193 | 17.1 |
| Critical care units | 261 | 23.1 |
| Infectious Disease / Isolation wards | 43 | 3.8 |
| General wards | 297 | 26.4 |
| OPD | 333 | 29.5 |

Of the respondents, 62.1% were working in tertiary institutions, while the rest split equally between primary and secondary centers. Regarding their clinical role, 42.7% were nurses, 39.1% physicians, and their clinical assignment area distributed fairly across the different departments of the healthcare institutions (Table 1).

Considering their COVID-19 status, 41.6% had COVID-19. About half of them travelled to countries that recently reported Monkeypox disease, mainly Europe, North America, and Australia (52.4%), UAE (23.3%) while only (5.8%) travelled to Western or Central Africa (Table 2). 60% of the surveyed HCWs’ rated their awareness level about the Monkeypox disease at the current time as moderate to high. While their self-rated worry level from the Monkeypox disease developing into a worldwide pandemic ranged from none/a little worried (48.7%), moderate (26%), to a lot worried (25.3%) (Table 2). (20.4%) of the HCWs perceived no worry at all travelling abroad considering the current Monkeypox outbreak, 66.1% were somewhat worried, and 13.5% were extremely worried. The main sources of worry a were acquiring the infection themselves or their families (49.6%), development of another worldwide pandemic (68.8%), or national lockdown (42.6%) and worry from international travel restrictions (40.8%) (Figure 1). When asked whether Monkeypox disease can cause more severe symptoms compared to smallpox disease, 23.4% disagreed, while the rest were split equally between agree or unsure.

**Table-2:** Descriptive analysis of the HCW’s Monkeypox disease perceptions and COVID-19 status.

| Variable | Frequency | Percentage |
| --- | --- | --- |
| **Have you been previously diagnosed with COVID19?** |  |  |
| Yes | 469 | 41.6 |
| No | 658 | 58.4 |
| **Have you travelled in the last month to a country where Monkeypox was recently reported?** |  |  |
| West or Central Africa | 7 | 5.8 |
| Europe, North America and Australia | 35 | 52.4 |
| UAE | 28 | 23.3 |
| Other (far Asia, India, Spain, France, and middle eastern countries) | 57 | 47.5 |
| **How would you rate your awareness about Monkeypox at the meantime? mean (SD)** |  |  |
| Low | 451 | 40.1 |
| Moderate | 415 | 36.8 |
| High | 261 | 23.2 |
| **How worried are you that Monkeypox can cause worldwide pandemic similar to COVID-19?** |  |  |
| None/little worried | 549 | 48.7 |
| Moderate worry | 293 | 26 |
| Worried a lot | 285 | 25.3 |
| **Do you think Monkeypox causes more severe disease compared to Smallpox?** |  |  |
| Disagree | 264 | 23.4 |
| Unsure | 448 | 39.8 |
| Agree | 415 | 36.8 |
| **Which is more worrisome to you, COVID-19 or Monkeypox disease?** |  |  |
| Unsure / Equally worried | 88 | 7.8 |
| I am more worried about COVID-19 | 637 | 56.5 |
| I am more worried about Monkeypox | 402 | 35.7 |
| **Healthcare workers should apply more infection control measures than the current ones, With the new Monkeypox outbreaks in some countries?** |  |  |
| Agree | 762 | 67.6 |
| Neither agree nor disagree | 292 | 25.9 |
| Disagree | 73 | 6.5 |
| **Please rate your worry level about traveling abroad with the new Monkeypox outbreaks in some countries** |  |  |
| Not worried at all | 230 | 20.4 |
| Somewhat worried | 745 | 66.1 |
| Extremely worried | 152 | 13.5 |
| **After Receiving this survey, did you perceive need to read more about Monkeypox disease?** |  |  |
| No | 291 | 25.8 |
| Yes | 836 | 74.2 |
| **Your sources of Information about Monkeypox disease** |  |  |
| Official local statements or press release from MOH or Saudi-CDC (Weqayah) | 608 | 57.6 |
| International health authorities' websites (WHO or CDC) | 631 | 59.8 |
| Social Networks (like YouTube, Facebook, Twitter, WhatsApp) | 539 | 51.1 |
| Scientific journals | 259 | 24.5 |
| Other Internet based sources | 32 | 3 |

**Figure 1:**
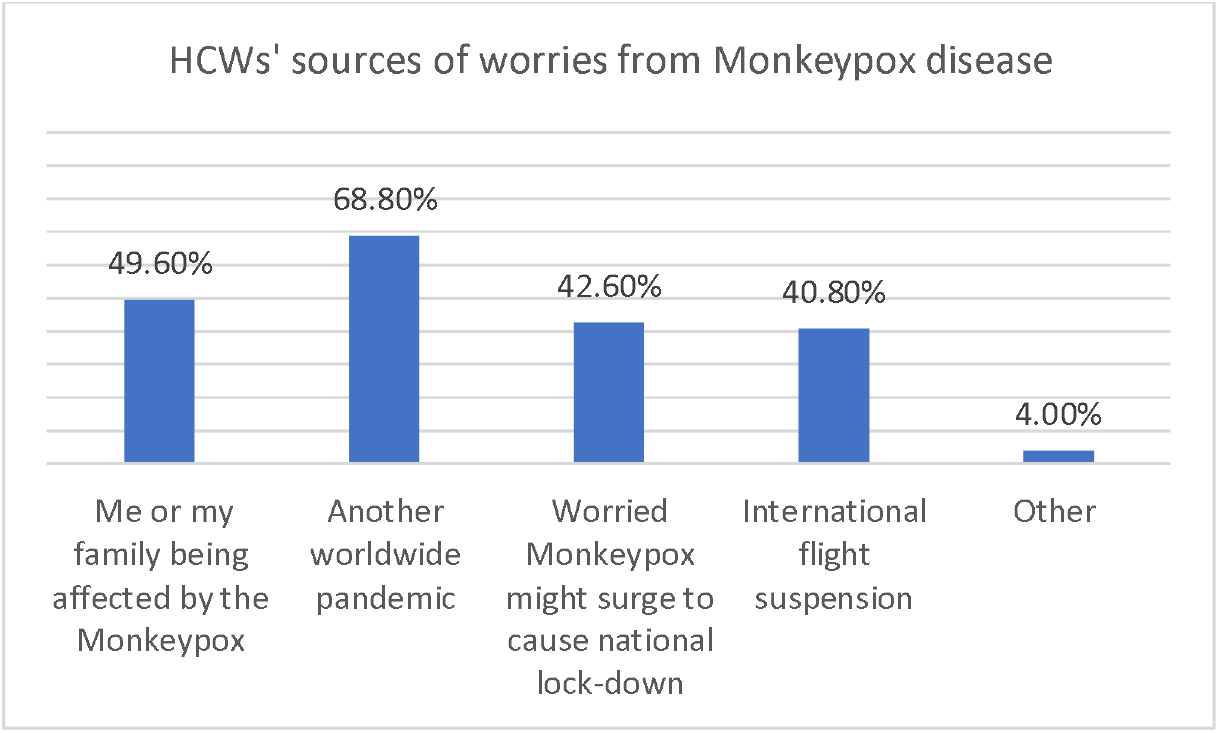
HCWs’ sources of worries from Monkeypox disease

Currently 56.5% of the HCW’s were more worried from COVID19 compared to Monkeypox, 35.7% of them were indeed more worried from Monkeypox, while 7.8% of the HCW’s were unsure or equally worried from both diseases. In relation to the need to implement more infection control measures than current ones considering Monkeypox outbreak, 67.6% agreed, 25.9% were unsure and 6.5% disagreed.

Interestingly, 74.2% of the surveyed HCWs perceived that they need to read more about the Monkeypox disease after receiving the current survey. Their sources of information about Monkeypox disease are detailed in Table 2.

Figure 2 shows the details of the HCWs’ HCWs prioritization for receptions of Monkeypox vaccine, 69.8% perceived that HCWs showed be vaccinated, followed by patients with immune deficiency 54.3%, then elderly 53.1%, then international travelers 40.4%.

**Figure 2:**
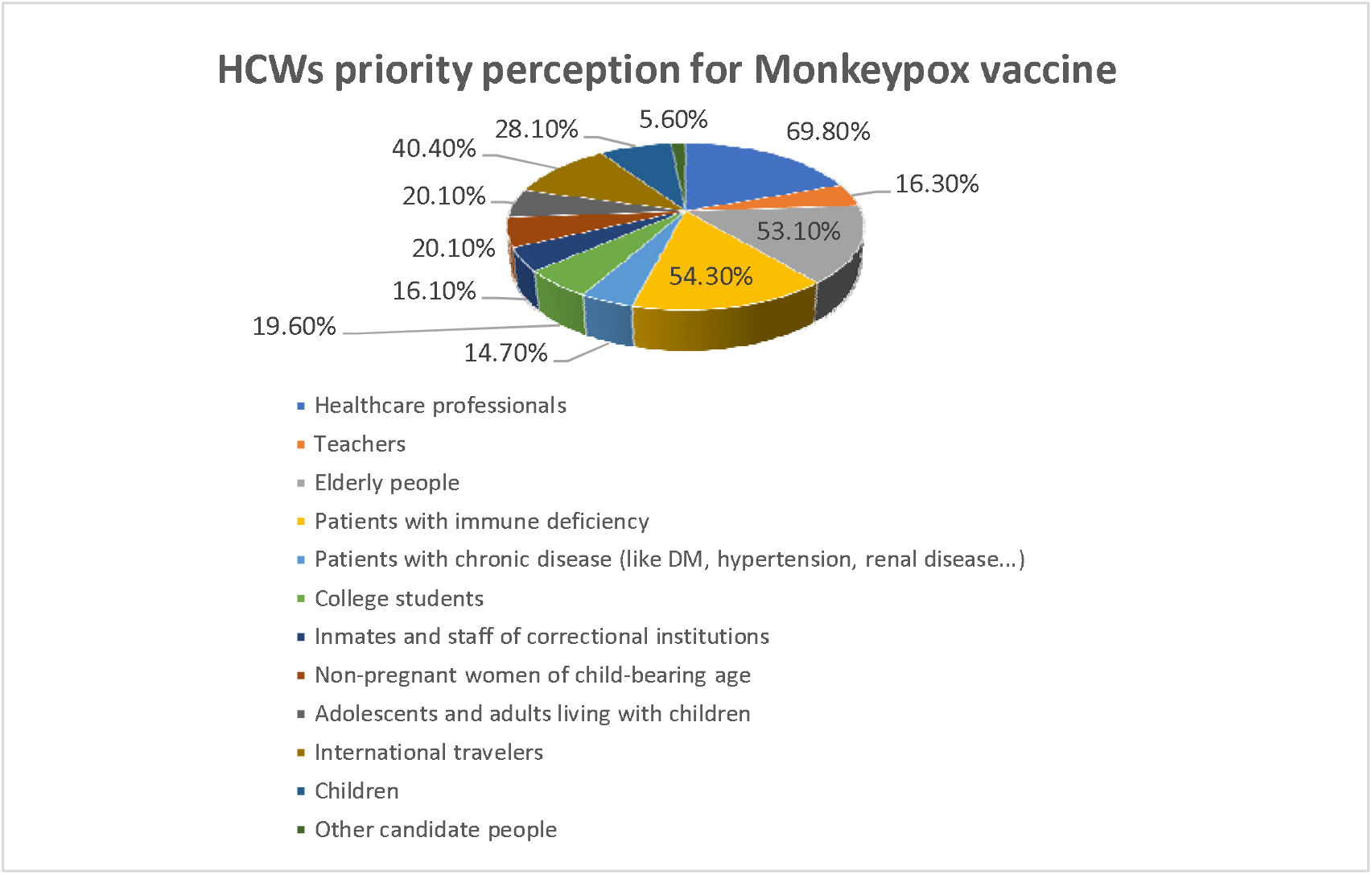
HCWs priority perception for Monkeypox vaccine

We assessed variables that were associated with HCW’s odds of having high worry from Monkeypox using multivariate Binary Logistic regression analysis Table 3. Males were significantly less predicted to have high worry from monkeypox compared to females (OR 0.533, p<0.001). While HCWs’ age, household’s size and GAD7 score did not correlate significantly with their odds of high worry from Monkeypox disease. Those who previously developed COVID-19 were significantly less worried (OR .735, p value .026). On the other side the participants’ clinical role showed that medical students were significantly more worried compared to the other surveyed participants (OR 2.79, p<0.001). HCWs’ self-rated awareness level of the Monkeypox disease correlated significantly but negatively with their odds of high worry from Monkeypox disease (OR=0.765, p <0.001). The HCWs’ worry from Monkeypox progression into pandemic correlated significantly and positively with their odds of high worry from the disease (OR1.465 p<0.001), in addition to their perception of Monkeypox as severe disease (OR 1.26, p-value 0.017) and those who agreed with application of tighter infection control measures of the current applied ones to control the disease (OR=1.691, p<0.001). While the HCWs’ worry from international travel restrictions did not correlate significantly with their odds of high worry from the Monkeypox disease p=0.151.

**Table-3:** Multivariate Binary Logistic Regression Analysis of the HCW’s odds of high worry from Monkeypox compared to COVID-19.

|  | **Multivariate**<br>**adjusted Odds Ratio**<br>**(OR)** | **95% CI**<br>**Lower** | **Upper** | **P value** |
| --- | --- | --- | --- | --- |
| Sex= Male | .533 | .394 | .721 | <0.001 |
| Age (years) | 1.000 | .984 | 1.017 | .964 |
| Previous COVID-19 infection | .735 | .560 | .964 | .026 |
| Households Size-Family size | 1.115 | .945 | 1.314 | .196 |
| Clinical Role<br>Medical student | 2.796 | 1.570 | 4.978 | <0.001 |
| Self-rated awareness of Monkeypox disease score | .765 | .661 | .885 | <0.001 |
| Worry from Monkeypox progression into pandemic | 1.465 | 1.275 | 1.682 | <0.001 |
| Perception of Monkeypox as severe disease | 1.263 | 1.043 | 1.529 | .017 |
| GAD7 score. | .987 | .958 | 1.017 | .393 |
| Agreement with application of tighter infection control measures | 1.691 | 1.294 | 2.210 | <0.001 |
| High worry from international travel restrictions | 1.206 | .934 | 1.557 | .151 |
| Constant | .056 |  |  | <0.001 |

**DV= High worry from Monkeypox disease**

Table-4 displays multivariate binary logistic regression analysis of the HCWs’ odds of agreement with vaccination of HCWs against the Monkeypox disease in the current stage of the outbreak. The HCWs’ sex, age, marital state, and healthcare institution type did not correlate significantly with their odds of agreement with vaccination. While those who developed COVID-19 previously were significantly more predicted to agree (OR 1.327, p-value=0.043). Also, the HCWs who supported application of tighter infection control measures compared to the current ones to combat Monkeypox disease were significantly more predicted to agree with vaccination (OR 1.415, p 0.003). The HCWs’ perception of need to read more about the disease and their GAD7 score did not correlate significantly with their odds of supporting vaccination. The HCWs’ sources of information correlated significantly and positively with their odds to support vaccination but with variable strength, those who relied on local official sources (OR 2.023, p<0.001) followed by social networks (OR 1.953, p<0.001) then international health websites (OR 1.375, p<0.001).

**Table 4:** Multivariate Binary Logistic Regression Analysis of the HCW’s odds of supporting vaccinations against Monkeypox disease.

|  | Multivariate<br>adjusted Odds<br>Ratio (OR) | 95% CI |  | p-value |
| --- | --- | --- | --- | --- |
|  |  | Lower | Upper |  |
| Sex | .899 | .670 | 1.206 | .478 |
| Male |  |  |  |  |
| Age (years) | .993 | .976 | 1.010 | .416 |
| Marital state | .891 | .629 | 1.261 | .515 |
| Ever married |  |  |  |  |
| Type of institution | 1.232 | .934 | 1.624 | .140 |
| Tertiary |  |  |  |  |
| Previous COVID-19 infection | 1.327 | 1.009 | 1.745 | .043 |
| Agreement with application of tighter infection control measures | 1.415 | 1.125 | 1.781 | .003 |
| HHI | 1.019 | .931 | 1.116 | .682 |
| Perception need to read more about Monkeypox | 1.248 | .896 | 1.737 | .189 |
| GAD7 | 1.006 | .977 | 1.037 | .673 |
| Source of information |  |  |  |  |
| Local official statements | 2.023 | 1.515 | 2.702 | <0.001 |
| International health | 1.375 | 1.031 | 1.833 | .030 |
| Social media channels | 1.953 | 1.468 | 2.598 | <0.001 |
| Constant | .054 |  |  | <0.001 |

**DV= HCW's odds of supporting vaccinations against Monkeypox disease.**

To further assess HCWs’ perception about the Monkeypox disease and their worries, we ran multivariate Binary Logistic regression analysis to assess the variables that are independently associated with their support to apply tighter infection control measures against Monkeypox disease compared to the currently applied ones after the COVID-19 pandemic Table 5. HCWs’ sex, age marital state, clinical role as well as their GAD7 score did not correlate significantly with their odds to support tighter measures.

**Table-5:** Multivariate Binary Logistic Regression Analysis of the HCW’s odds of supporting Implementation of tighter infection control measures against Monkeypox compared to the currently applied.

|  | Multivariate<br>adjusted<br>Odds Ratio<br>(OR) | 95% CI |  | p-value |
| --- | --- | --- | --- | --- |
|  |  | Lower | Upper |  |
| Sex | .863 | .631 | 1.179 | .354 |
| Male |  |  |  |  |
| Age (years) | .982 | .964 | 1.001 | .060 |
| Ever married | 1.365 | .936 | 1.991 | .106 |
| Clinical role | .891 | .743 | 1.069 | .213 |
| Perception to read more about Monkeypox | 3.157 | 2.268 | 4.393 | <0.001 |
| GAD7 | 1.012 | .978 | 1.048 | .490 |
| Source Of information |  |  |  |  |
| International health websites | 1.242 | .900 | 1.712 | .187 |
| Social media /networks channels | 1.353 | 1.005 | 1.820 | .046 |
| Science journals | 0.656 | .456 | 0.943 | .023 |
| More worried from Monkeypox compared to COVID-19 | 2.067 | 1.469 | 2.908 | <0.001 |
| Worry level from international travel restrictions | 1.568 | 1.184 | 2.078 | .002 |
| Household size>=4 persons | 1.230 | 1.031 | 1.466 | .021 |
| Worry from Monkeypox progress into pandemic | 0.813 | 0.703 | 0.941 | .006 |
| Perception of Monkeypox as severe disease | 1.316 | 1.072 | 1.617 | .009 |
| Constant | 0.120 |  |  | .002 |

**DV= Supporting implementation of tighter infection control measures against Monkeypox compared to the currently applied**

However, their perceived worry from the severity of the Monkeypox disease correlated significantly and positively with their odds of supporting the application of tighter infection control measures against the monkeypox disease, (OR 1.316, p .009). This was reflected also on their perception of need to read more about Monkeypox, which was associated significantly and positively with support to apply tighter measures (OR 3.157, p<0.001). Considering their sources of information about Monkeypox disease, use of social media networks was associated with high support (OR 1.35, p .046), while those who relied mainly on scientific journal were less likely to support application of tighter infection control measures (OR 0.656, p .023), and those who relied on international health websites did not have any convergence with their odds to support tighter measures.

HCWs who were more worried from Monkeypox compared to COVID-19 were significantly more predicted to support implementing tighter infection control measures compared to currently applied ones (OR 2.067, p<0.001). Regarding HCWs’ sources of worry from Monkeypox disease, those whom their main worry was international travel restrictions had higher support for the application of tighter measures (OR 1.568, p .002), while those who were mainly worried about Monkeypox outbreak progression into a pandemic had significantly lower support to apply tighter measures (OR 0.813, p .006). HCWs’ household size correlated with their odds of supporting the implementation of tighter infection control measures, participants living with > 3 persons were significantly more predicted to support application of tighter infection control measures (OR 1.230, p=0.021).

## Discussion

The international health care system experienced one of the largest and most stressful pandemics in the recent decades, namely the COVID-19, which has put tremendous burden. This was related to expansion demand within very short time and high levels of anxiety and worry of acquiring the disease. With the recovery of the health care system after those two years of pandemic, and massive vaccination campaigns that targeted not only medical practitioners but also public, this demanded and put extra efforts on the health care system. The reemergence of the Monkeypox disease just within the resolution phase of the COVID-19 pandemic has been a challenging alert for the health care system especially with loosing of the already applied infection prevention measures during the COVID-19 pandemic. The Monkeypox disease has been described the first time in 1970 in the Democratic Republic of the Congo[20]. Since then, multiple cases have been described mainly in African countries with occasional outbreaks as in 1996–97 in the Democratic Republic of the Congo and Nigeria in 2017[20]. Still, it has been described outside Africa since 2003. The recent re-emergence of Monkeypox in outbreak fashion in multiple countries simultaneously and its relation to sexual activity and intimate body interaction, such as men having sex with men (MSM), participating in extended sexual networks, and the multiple modes of transmission of the disease, involving respiratory droplets, skin lesions, and sexual contact, all heightened the alert within the medical system[21].

Our study has shown that 56.5% of the HCWs were more worried from COVID19 compared to Monkeypox, 48.7% had no to a little worry about its progression into a worldwide pandemic while 25.3% were a lot worried. 67.6% were in agreement of applying tighter infection control measure compared to the currently applied once in order to combat the current outbreak of Monkeypox disease.

Furthermore, HCWs’ self-rated awareness level of the Monkeypox disease, expectedly, correlated significantly but negatively with their odds of high worry from Monkeypox disease. Similarly, the HCWs’ worry from Monkeypox progress into pandemic correlates with their perceived severity of the Monkeypox disease and agreement with tighter infection control measures.

Male HCWs within Saudi Arabia were less predicted to worry about Monkeypox compared to females, which is a common observation of gender difference in relation to infectious disease risk such as COVID-19 in HCWs[22,23]. This difference, interestingly, resulted in huge mental health burden and impact on workforce[24]. On the other side, the participants’ clinical role showed that medical students were significantly more worried compared to the other surveyed participants. This finding was not surprising as COVID-19 pandemic affected medical students more and even raised their concerns to reconsider their profession[25]. HCWs who previously developed COVID-19 were also significantly less worried about the Monkeypox disease outbreak, most probably that is related to surviving already through the COVID-19 pandemic and even the infection itself, some reports have shown shorter recovery time from COVID-19 and milder symptoms in HCWs, that and their medical knowledge background of the Monkeypox disease with its limited potential of transmission compared to COVID-19, mostly led to lower worry level in HCWs COVID-19 survivors[26]. In line with the HCWs’ medical knowledge background about the Monkeypox disease, its vulnerability, and their ability to critically interpret the transmission facts about its most recent outbreak, their self-rated awareness of the disease correlated negatively with their odds of high worry level from the disease. While HCWs who had high concern of Monkeypox progression into pandemic and those who perceived the disease potentially being severe had high worry level. Similar findings were also observed for those potentially perceived the disease as a threat nevertheless the scientific background basis of their perception, similar observation has been described in HCWs in contagion with COVID-19 patients[27,28]. In line with the previous observation, HCWs who perceived need to apply tighter infection control measures of the currently applied to limit Monkeypox progress also had odds of high worry from the disease. Such observations of HCWs’ perceptions of Monkeypox disease, its severity and how to combat it are similar to COVID-19 studies and might reflect the anxious temperament of the respondents[29].

Currently there are two vaccines that are being suggested against Monkeypox. ACAM2000 is a second-generation smallpox vaccine made from vaccinia virus and has activity against Monkeypox[30], and JYNNEOS vaccine (also known as Imvanex, or Imvamune) is a live non-replicating vaccine produced from a modified strain of Vaccinia Ankara-Bavarian Nordic (MVA-BN) orthopoxvirus, which has been previously given to HCWs who were involved in investigating Monkeypox outbreaks[31]. Advocacy for vaccinating HCWs against Monkeypox deserves further studies in the context of reemergence of the disease and recent outbreaks. The current study did not find any correlation between HCWs different demographics and vaccine acceptance, however, those who had been previously infected with COVID-19 were more likely to support vaccination against Monkeypox. A previous study from KSA that examined factors related to COVID-19 vaccine acceptance among HCWs prior to its availability did not show higher acceptance among previously infected individuals with COVID-19, but probably after 2 years of the pandemic and experiencing the protection effect of vaccination campaigns, HCWs felt more encouraged toward Monkeypox vaccination with increasing confidence in vaccination in general, this was observed with Flu vaccine during COVID-19 pandemic and vaccination in general[32-34]. Interestingly, HCWs who advocated for tighter infection-prevention and control measures were also more supportive of vaccination, both in line with their healthy behavior and their belief that both are integral to each other, this echoes a previous study that showed higher rate of hepatitis B vaccination in HCWs who attended infection control training[35].

Other possible infection prevention and control measures that may be applicable if the outbreak evolves further may include preexposure prophylaxis with the vaccines[36]. This seems to be welcomed by the surveyed HCWs. Furthermore, we identified that HCWs in KSA who rely on official local statements from Saudi MOH and CDC were more likely to accept vaccination. Reliance on the latter sources have been previously proven to increase COVID-19 vaccine acceptance in numerous studies from KSA, this study further supports utilizing these sources for accurate and reliable information[15,17,37,38].

In this study, there was a significant correlation between high level of worry from Monkeypox disease and the need to expand HCWs’ information about the disease from one side and support to have tighter infection control measures. It is interesting to note that relying on medical and scientific journal was not associated with HCWs’ believe in tighter infection control measures to combat the outbreak, this was in contradiction to those who relied on social media. That is in line with previous studies of COVID-19 pandemic among HCWs which showed that the top three sources of HCWs for information were social media, WHO website, and Saudi ministry of health[17,38,39]. It had been noted that relying on official and reliable sources for information about pandemics had enhanced the perceptions of HCWs’ about the pandemics and vaccination[15].

HCWs with increased worry about the emergence of multi-country outbreak of Monkeypox disease, and worry of international travel restrictions were predicted to support implementing tighter infection control measures compared to currently applied measures. This worry could be further fueled by the recent announcement of the World Health Organization (WHO) that the current outbreak constitutes a public health emergency of international concern (PHEIC)[40]. In addition, HCWs with a family size of > 3 persons were significantly predicted to support application of tighter infection control measures. The family size would heighten alert and put high burden as well as increase odd for hospitalization as shown in one study that family size of 4 or more which was 2.5[41].

### Strengths and limitations

The current study carries the advantage of earlier exploration of HCWs on the perceptions, worries and vaccination support against evolving Monkeypox disease within the first month of the WHO alert of its reemergence as outbreaks in multiple countries outside the African continent. This current alert by the WHO is worth studying due to the exceptional demand faced by the fatiguing international healthcare system by the resolving COVID-19 pandemic. Still, the current study has limitations related to its survey-based design, such as the convenience sampling technique and recall bias. Another limitation is related to scarcity of such cases in the Saudi healthcare system and unfamiliarity of HCWs with the disease, which may affect their perceptions, worry level and even knowledge. Even so, the current study gives highlight to local healthcare administratives about the HCWs’ perceptions and readiness for Monkeypox especially in case it progresses into local or international pandemic.

## Conclusions

The current study has shown that almost half of HCWs within Saudi Arabia are more worried about Monkeypox disease compared to COVID-19 and its potential progression into another pandemic. In addition, the majority were in favor of applying tighter infection prevention measures to combat the disease. The current study highlights areas needed for the healthcare administrative about the HCWs’ perceptions and readiness for Monkeypox especially in the event of the occurrence of local or international pandemic.

## Data Availability

All data produced in the present study are available upon reasonable request to the corresponding author.

## Abbreviations

CDC: Centers for Disease Control and Prevention
COVID-19: Coronavirus disease 2019
MOH: Ministry of Health
MOXV: Monkeypox virus
SARS-CoV-2: Severe acute respiratory syndrome coronavirus 2
WHO: World Health Organization

## Acknowledgments

The authors extend their appreciation to the Deanship of Scientific Research, King Saud University, for funding through Vice Deanship of Scientific Research Chairs; Research Chair of Prince Abdullah Ben Khalid Celiac Disease; Riyadh, Kingdom of Saudi Arabia. The research team is thankful for the statistical data analysis consultation offered by www.hodhodata.com

## Refs

1. Al-Tawfiq, J.A.; Barry, M.; Memish, Z.A. International outbreaks of Monkeypox virus infection with no established travel: A public health concern with significant knowledge gap. Travel Med Infect Dis 2022, 49, 102364, DOI:10.1016/j.tmaid.2022.102364.

2. León-Figueroa, D.A.; Bonilla-Aldana, D.K.; Pachar, M.; Romaní, L.; Saldaña-Cumpa, H.M.; Anchay-Zuloeta, C.; Diaz-Torres, M.; Franco-Paredes, C.; Suárez, J.A.; Ramirez, J.D.; et al. The never-ending global emergence of viral zoonoses after COVID-19? The rising concern of monkeypox in Europe, North America and beyond. Travel Med Infect Dis 2022, 49, 102362, DOI:10.1016/j.tmaid.2022.102362.

3. Kozlov, M. Monkeypox goes global: why scientists are on alert. In Nature; England, 2022; Volume 606, pp. 15–16.

4. WHO. Meeting of the International Health Regulations (2005) Emergency Committee regarding the multi-country monkeypox outbreak. Available online: https://www.who.int/news/item/25-06-2022-meeting-of-the-international-health-regulations-(2005)-emergency-committee--regarding-the-multi-country-monkeypox-outbreak (accessed on 1 Aug 2022).

5. Temsah, M.-H.; Aljamaan, F.; Alenezi, S.; Alhasan, K.; Saddik, B.; Al-Barag, A.; Alhaboob, A.; Bahabri, N.; Alshahrani, F.; Alrabiaah, A.; et al. Monkeypox caused less worry than COVID-19 among the general population during the first month of the WHO Monkeypox alert. medRxiv 2022, 2022.2007.2007.22277365, DOI:10.1101/2022.07.07.22277365.

6. Al-Tawfiq, J.A.; Temsah, M.H. Perspective on the challenges of COVID-19 facing healthcare workers. In Infection; © 2022. The Author(s), under exclusive licence to Springer-Verlag GmbH Germany.: Germany, 022.

7. Kozlov, M. Monkeypox vaccination begins - can the global outbreaks be contained? Nature 2022, 606, 444–445, DOI:10.1038/d41586-022-01587-1.

8. Selb, R.; Werber, D.; Falkenhorst, G.; Steffen, G.; Lachmann, R.; Ruscher, C.; McFarland, S.; Bartel, A.; Hemmers, L.; Koppe, U.; et al. A shift from travel-associated cases to autochthonous transmission with Berlin as epicentre of the monkeypox outbreak in Germany, May to June 2022. Euro Surveill 2022, 27, DOI:10.2807/1560-7917.es.2022.27.27.2200499.

9. Iñigo Martínez, J.; Gil Montalbán, E.; Jiménez Bueno, S.; Martín Martínez, F.; Nieto Juliá, A.; Sánchez Díaz, J.; García Marín, N.; Córdoba Deorador, E.; Nunziata Forte, A.; Alonso García, M.; et al. Monkeypox outbreak predominantly affecting men who have sex with men, Madrid, Spain, 26 April to 16 June 2022. Euro Surveill 2022, 27, DOI:10.2807/1560-7917.es.2022.27.27.2200471.

10. McAndrew, T.; Majumder, M.S.; Lover, A.A.; Venkatramanan, S.; Bocchini, P.; Besiroglu, T.; Codi, A.; Braun, D.; Dempsey, G.; Abbott, S.; et al. Early human judgment forecasts of human monkeypox, May 2022. Lancet Digit Health 2022, 4, e569–e571, DOI:10.1016/s2589-7500(22)00127-3.

11. Organization, W.H. Vaccines and immunization for monkeypox: interim guidance, 14 June 2022; World Health Organization: 2022.

12. Simpson, L.A.; Macdonald, K.; Searle, E.F.; Shearer, J.A.; Dimitrov, D.; Foley, D.; Morales, E.; Shenoy, E.S. Development and deployment of tools for rapid response notification of Monkeypox exposure, exposure risk assessment and stratification, and symptom monitoring. Infect Control Hosp Epidemiol 2022, 1–5, DOI:10.1017/ice.2022.167.

13. Temsah, M.H.; Alhuzaimi, A.N.; Alamro, N.; Alrabiaah, A.; AlSohime, F.; Alhasan, K.; Kari, J.A.; Almaghlouth, I.; Aljamaan, F.; Al Amri, M.; et al. Knowledge, Attitudes, and Practices of Healthcare Workers During the Early COVID-19 Pandemic in a Main, Academic Tertiary Care Centre in Saudi Arabia. Epidemiol Infect 2020, 1–29, DOI:10.1017/S0950268820001958.

14. Temsah, M.H.; Al-Sohime, F.; Alamro, N.; Al-Eyadhy, A.; Al-Hasan, K.; Jamal, A.; Al-Maglouth, I.; Aljamaan, F.; Al Amri, M.; Barry, M.; et al. The psychological impact of COVID-19 pandemic on health care workers in a MERS-CoV endemic country. J Infect Public Health 2020, 13, 877–882, DOI:10.1016/j.jiph.2020.05.021.

15. Temsah, M.H.; Barry, M.; Aljamaan, F.; Alhuzaimi, A.N.; Al-Eyadhy, A.; Saddik, B.; Alsohime, F.; Alhaboob, A.; Alhasan, K.; Alaraj, A.; et al. SARS-CoV-2 B.1.1.7 UK Variant of Concern Lineage-Related Perceptions, COVID-19 Vaccine Acceptance and Travel Worry Among Healthcare Workers. Front Public Health 2021, 9, 686958, DOI:10.3389/fpubh.2021.686958.

16. Barry, M.; Temsah, M.-H.; Aljamaan, F.; Saddik, B.; Al-Eyadhy, A.; Alanazi, S.; Alamro, N.; Alhuzaimi, A.; Alhaboob, A.; Alsohime, F. COVID-19 vaccine uptake among healthcare workers in the fourth country to authorize BNT162b2 during the first month of rollout. medRxiv 2021.

17. Alhasan, K.; Aljamaan, F.; Temsah, M.H.; Alshahrani, F.; Bassrawi, R.; Alhaboob, A.; Assiri, R.; Alenezi, S.; Alaraj, A.; Alhomoudi, R.I.; et al. COVID-19 Delta Variant: Perceptions, Worries, and Vaccine-Booster Acceptability among Healthcare Workers. 2Healthcare (Basel) 2021, 9, DOI:10.3390/healthcare9111566.

18. Spitzer, R.L.; Kroenke, K.; Williams, J.B.; Lowe, B. A brief measure for assessing generalized anxiety disorder: the GAD-7. 2Arch Intern Med 2006, 166, 1092–1097, DOI:10.1001/archinte.166.10.1092.

19. AlHadi, A.N.; AlAteeq, D.A.; Al-Sharif, E.; Bawazeer, H.M.; Alanazi, H.; AlShomrani, A.T.; Shuqdar, R.M.; AlOwaybil, R. An arabic translation, reliability, and validation of Patient Health Questionnaire in a Saudi sample. Ann Gen Psychiatry 2017, 16, 32, DOI:10.1186/s12991-017-0155-1.

20. WHO. Monkeypox. Available online: https://www.who.int/news-room/fact-sheets/detail/monkeypox (accessed on 1 Aug 2022).

21. Petersen, E.; Abubakar, I.; Ihekweazu, C.; Heymann, D.; Ntoumi, F.; Blumberg, L.; Asogun, D.; Mukonka, V.; Lule, S.A.; Bates, M.; et al. Monkeypox - Enhancing public health preparedness for an emerging lethal human zoonotic epidemic threat in the wake of the smallpox post-eradication era. Int J Infect Dis 2019, 78, 78–84, DOI:10.1016/j.ijid.2018.11.008.

22. Spoorthy, M.S.; Pratapa, S.K.; Mahant, S. Mental health problems faced by healthcare workers due to the COVID-19 pandemic-A review. Asian J Psychiatr 2020, 51, 102119, DOI:10.1016/j.ajp.2020.102119.

23. Huang, Q.; Luo, L.S.; Wang, Y.Y.; Jin, Y.H.; Zeng, X.T. Gender Differences in Psychological and Behavioral Responses of Infected and Uninfected Health-Care Workers During the Early COVID-19 Outbreak. Front Public Health 2021, 9, 638975, DOI:10.3389/fpubh.2021.638975.

24. Liu, S.; Yang, L.; Zhang, C.; Xu, Y.; Cai, L.; Ma, S.; Wang, Y.; Cai, Z.; Du, H.; Li, R.; et al. Gender differences in mental health problems of healthcare workers during the coronavirus disease 2019 outbreak. J Psychiatr Res 2021, 137, 393–400, DOI:10.1016/j.jpsychires.2021.03.014.

25. Gupta, P.; B, K.A.; Ramakrishna, K. Prevalence of Depression and Anxiety Among Medical Students and House Staff During the COVID-19 Health-Care Crisis. Acad Psychiatry 2021, 45, 575–580, DOI:10.1007/s40596-021-01454-7.

26. Breugnon, E.; Thollot, H.; Fraissenon, A.; Saunier, F.; Labetoulle, R.; Pillet, S.; Lucht, F.; Berthelot, P.; Botelho-Nevers, E.; Gagneux-Brunon, A. COVID-19 outpatient management: Shorter time to recovery in Healthcare workers according to an electronic daily symptoms assessment. Infect Dis Now 2021, 51, 71–76, DOI:10.1016/j.medmal.2020.10.001.

27. Trumello, C.; Bramanti, S.M.; Ballarotto, G.; Candelori, C.; Cerniglia, L.; Cimino, S.; Crudele, M.; Lombardi, L.; Pignataro, S.; Viceconti, M.L.; et al. Psychological Adjustment of Healthcare Workers in Italy during the COVID-19 Pandemic: Differences in Stress, Anxiety, Depression, Burnout, Secondary Trauma, and Compassion Satisfaction between Frontline and Non-Frontline Professionals. Int J Environ Res Public Health 2020, 17, DOI:10.3390/ijerph17228358.

28. Buselli, R.; Corsi, M.; Baldanzi, S.; Chiumiento, M.; Del Lupo, E.; Dell’Oste, V.; Bertelloni, C.A.; Massimetti, G.; Dell’Osso, L.; Cristaudo, A.; et al. Professional Quality of Life and Mental Health Outcomes among Health Care Workers Exposed to Sars-Cov-2 (Covid-19). Int J Environ Res Public Health 2020, 17, DOI:10.3390/ijerph17176180.

29. Moccia, L.; Janiri, D.; Pepe, M.; Dattoli, L.; Molinaro, M.; De Martin, V.; Chieffo, D.; Janiri, L.; Fiorillo, A.; Sani, G.; et al. Affective temperament, attachment style, and the psychological impact of the COVID-19 outbreak: an early report on the Italian general population. Brain Behav Immun 2020, 87, 75–79, DOI:10.1016/j.bbi.2020.04.048.

30. Russo, A.T.; Berhanu, A.; Bigger, C.B.; Prigge, J.; Silvera, P.M.; Grosenbach, D.W.; Hruby, D. Co-administration of tecovirimat and ACAM2000™ in non-human primates: Effect of tecovirimat treatment on ACAM2000 immunogenicity and efficacy versus lethal monkeypox virus challenge. Vaccine 2020, 38, 644–654, DOI:10.1016/j.vaccine.2019.10.049.

31. Costello, V.; Sowash, M.; Gaur, A.; Cardis, M.; Pasieka, H.; Wortmann, G.; Ramdeen, S. Imported Monkeypox from International Traveler, Maryland, USA, 2021. Emerg Infect Dis 2022, 28, 1002–1005, DOI:10.3201/eid2805.220292.

32. Barry, M.; Temsah, M.H.; Alhuzaimi, A.; Alamro, N.; Al-Eyadhy, A.; Aljamaan, F.; Saddik, B.; Alhaboob, A.; Alsohime, F.; Alhasan, K.; et al. COVID-19 vaccine confidence and hesitancy among health care workers: A cross-sectional survey from a MERS-CoV experienced nation. PLoS One 2021, 16, e0244415, DOI:10.1371/journal.pone.0244415.

33. Bertoni, L.; Roncadori, A.; Gentili, N.; Danesi, V.; Massa, I.; Nanni, O.; Altini, M.; Gabutti, G.; Montella, M.T. How has COVID-19 pandemic changed flu vaccination attitudes among an Italian cancer center healthcare workers? Hum Vaccin Immunother 2022, 18, 1978795, DOI:10.1080/21645515.2021.1978795.

34. Pérez-Rivas, F.J.; Gallego-Lastra, R.D.; Marques-Vieira, C.M.A.; López-López, C.; Domínguez-Fernández, S.; Rico-Blázquez, M.; Ajejas Bazán, M.J. The Attitude towards Vaccination of Health Sciences Students at a Spanish University Improved over the First 18 Months of the COVID-19 Pandemic. Vaccines (Basel) 2022, 10, DOI:10.3390/vaccines10020237.

35. Akibu, M.; Nurgi, S.; Tadese, M.; Tsega, W.D. Attitude and Vaccination Status of Healthcare Workers against Hepatitis B Infection in a Teaching Hospital, Ethiopia. Scientifica (Cairo) 2018, 2018, 6705305, DOI:10.1155/2018/6705305.

36. Rao, A.K.; Petersen, B.W.; Whitehill, F.; Razeq, J.H.; Isaacs, S.N.; Merchlinsky, M.J.; Campos-Outcalt, D.; Morgan, R.L.; Damon, I.; Sánchez, P.J.; et al. Use of JYNNEOS (Smallpox and Monkeypox Vaccine, Live, Nonreplicating) for Preexposure Vaccination of Persons at Risk for Occupational Exposure to Orthopoxviruses: Recommendations of the Advisory Committee on Immunization Practices - United States, 2022. MMWR Morb Mortal Wkly Rep 2022, 71, 734–742, DOI:10.15585/mmwr.mm7122e1.

37. Temsah, M.H.; Aljamaan, F.; Alenezi, S.; Alhasan, K.; Alrabiaah, A.; Assiri, R.; Bassrawi, R.; Alhaboob, A.; Alshahrani, F.; Alarabi, M.; et al. SARS-CoV-2 Omicron Variant: Exploring Healthcare Workers’ Awareness and Perception of Vaccine Effectiveness: A National Survey During the First Week of WHO Variant Alert. Front Public Health 2022, 10, 878159, DOI:10.3389/fpubh.2022.878159.

38. Temsah, M.H.; Barry, M.; Aljamaan, F.; Alhuzaimi, A.; Al-Eyadhy, A.; Saddik, B.; Alrabiaah, A.; Alsohime, F.; Alhaboob, A.; Alhasan, K.; et al. Adenovirus and RNA-based COVID-19 vaccines’ perceptions and acceptance among healthcare workers in Saudi Arabia: a national survey. BMJ Open 2021, 11, e048586, DOI:10.1136/bmjopen-2020-048586.

39. Temsah, M.H.; Alenezi, S.; Alarabi, M.; Aljamaan, F.; Alhasan, K.; Assiri, R.; Bassrawi, R.; Alshahrani, F.; Alhaboob, A.; Alaraj, A.; et al. Healthcare Workers’ SARS-CoV-2 Omicron Variant Uncertainty-Related Stress, Resilience, and Coping Strategies during the First Week of the World Health Organization’s Alert. Int J Environ Res Public Health 2022, 19, DOI:10.3390/ijerph19041944.

40. WHO. Second meeting of the International Health Regulations (2005) (IHR) Emergency Committee regarding the multi-country outbreak of monkeypox. Available online: https://www.who.int/news/item/23-07-2022-second-meeting-of-the-international-health-regulations-(2005)-(ihr)-emergency-committee-regarding-the-multi-country-outbreak-of-monkeypox (accessed on 1 Aug 2022).

41. Nash, D.; Qasmieh, S.; Robertson, M.; Rane, M.; Zimba, R.; Kulkarni, S.G.; Berry, A.; You, W.; Mirzayi, C.; Westmoreland, D.; et al. Household factors and the risk of severe COVID-like illness early in the U.S. pandemic. PLoS One 2022, 17, e0271786, DOI:10.1371/journal.pone.0271786.

